# Feasibility of controlling 2019-nCoV outbreaks by isolation of cases and contacts

**DOI:** 10.1101/2020.02.08.20021162

**Authors:** Joel Hellewell, Sam Abbott, Amy Gimma, Nikos I Bosse, Christopher I Jarvis, Timothy W Russell, James D Munday, Adam J Kucharski, W John Edmunds, CMMID nCoV working group, Sebastian Funk, Rosalind M Eggo

## Abstract

**Background:** To assess the viability of isolation and contact tracing to control onwards transmission from imported cases of 2019-nCoV.

**Methods:** We developed a stochastic transmission model, parameterised to the 2019-nCoV outbreak. We used the model to quantify the potential effectiveness of contact tracing and isolation of cases at controlling a 2019 nCoV-like pathogen. We considered scenarios that varied in: the number of initial cases; the basic reproduction number *R*_*0*_; the delay from symptom onset to isolation; the probability contacts were traced; the proportion of transmission that occurred before symptom onset, and the proportion of subclinical infections. We assumed isolation prevented all further transmission in the model. Outbreaks were deemed controlled if transmission ended within 12 weeks or before 5000 cases in total. We measured the success of controlling outbreaks using isolation and contact tracing, and quantified the weekly maximum number of cases traced to measure feasibility of public health effort.

**Findings:** While simulated outbreaks starting with only 5 initial cases, *R*_*0*_ of 1.5 and little transmission before symptom onset could be controlled even with low contact tracing probability, the prospects of controlling an outbreak dramatically dropped with the number of initial cases, with higher *R*_*0*_, and with more transmission before symptom onset. Across different initial numbers of cases, the majority of scenarios with an R_0_ of 1.5 were controllable with under 50% of contacts successfully traced. For *R*_*0*_ of 2.5 and 3.5, more than 70% and 90% of contacts respectively had to be traced to control the majority of outbreaks. The delay between symptom onset and isolation played the largest role in determining whether an outbreak was controllable for lower values of *R*_*0*_. For higher values of *R*_*0*_ and a large initial number of cases, contact tracing and isolation was only potentially feasible when less than 1% of transmission occurred before symptom onset.

**Interpretation:** We found that in most scenarios contact tracing and case isolation alone is unlikely to control a new outbreak of 2019-nCov within three months. The probability of control decreases with longer delays from symptom onset to isolation, fewer cases ascertained by contact tracing, and increasing transmission before symptoms. This model can be modified to reflect updated transmission characteristics and more specific definitions of outbreak control to assess the potential success of local response efforts.

**Funding:** Wellcome Trust, Global Challenges Research Fund, and HDR UK.

**Research in Context:** *Evidence before this study:* Contact tracing and isolation of cases is a commonly used intervention for controlling infectious disease outbreaks. This intervention can be effective, but may require intensive public health effort and cooperation to effectively reach and monitor all contacts. When the pathogen has infectiousness before symptom onset, control of outbreaks using contact tracing and isolation is more challenging.

*Added value of this study:* This study uses a mathematical model to assess the feasibility of contact tracing and case isolation to control outbreaks of 2019-nCov, a newly emerged pathogen. We used disease transmission characteristics specific to the pathogen and therefore give the best available evidence if contact tracing and isolation can achieve control of outbreaks.

*Implications of all the available evidence:* Contact tracing and isolation may not contain outbreaks of 2019-nCoV unless very high levels of contact tracing are achieved. Even in this case, if there is asymptomatic transmission, or a high fraction of transmission before onset of symptoms, this strategy may not achieve control within three months.

## Introduction

As of 5th February 2020, there have been over 24,550 confirmed cases of a novel coronavirus infection (2019-nCoV), including over 190 international cases, and over 490 reported deaths^1^. Control measures have been instigated within China to try to contain the outbreak^2^. As infectious people arrive in countries or areas without ongoing transmission, efforts are being made to halt transmission, and prevent potential outbreaks^3,4^. Isolation of confirmed and suspected cases, and identification of contacts are a critical part of these control efforts. It is not yet clear if these efforts will achieve control of transmission of 2019- nCoV.

Isolation of cases and contact tracing becomes less effective if infectiousness begins before the onset of symptoms^5,6^. For example, the severe acute respiratory syndrome (SARS) outbreak that began in Southern China in 2003 was amenable to eventual control through tracing contacts of suspected cases and isolating confirmed cases because the majority of transmission occurred after symptom onset^7^. These interventions also play a major role in response to outbreaks where onset of symptoms and infectiousness are concurrent^10^, for example Ebola virus disease^8,9^ and MERS^10,11^, and for many other infections^12,13^.

The effectiveness of isolation and contact tracing methods hinges on two key epidemiological parameters: the number of secondary infections generated by each new infection and the proportion of transmission that occurs prior to symptom onset^5^. In addition, the probability of successful contact tracing and the delay between symptom onset and isolation are critical, since cases remain in the community where they can infect others until isolation^6,14^. Transmission prior to symptom onset could only be prevented by tracing contacts of confirmed cases and testing (and quarantining) those contacts. Cases that do not seek care, potentially due to subclinical or asymptomatic transmission represent a further challenge to control.

If 2019-nCoV can be controlled by isolation and contact tracing, then public health efforts should be focussed on this strategy. However, if this is not enough to control outbreaks, then additional resources may be needed for additional interventions. There are currently key unknown characteristics of the transmissibility and natural history of 2019-nCoV; for example, whether transmission can occur before symptom onset. Therefore we explored a range of epidemiological scenarios that represent potential transmission properties based on current information about 2019-nCoV transmission. We assessed the ability of isolation and contact tracing to control disease outbreaks using a mathematical model^6,15–18^. By varying the efficacy of contact tracing efforts, the size of the outbreak when detected, and the promptness of isolation after symptom onset, we show how viable it is for countries at risk of imported cases to use contact tracing and isolation as a containment strategy.

## Methods

### Model structure

We implemented a branching process model in which the number of potential secondary cases produced by each individual (the ‘infector’) is drawn from a negative binomial distribution with a mean equal to the reproduction number, and heterogeneity in the number of new infections produced by each individual^6,15,18–20^. Each potential new infection was assigned a time of infection drawn from the serial interval distribution. Secondary cases were only created if the infector had not been isolated by the time of infection. In the example in Figure 1, person A can potentially produce three secondary infections (because three is drawn from the negative binomial distribution), but only two transmissions occur before the case was isolated. Thus, a reduced delay from onset to isolation reduced the average number of secondary cases in the model.

**Figure 1:**
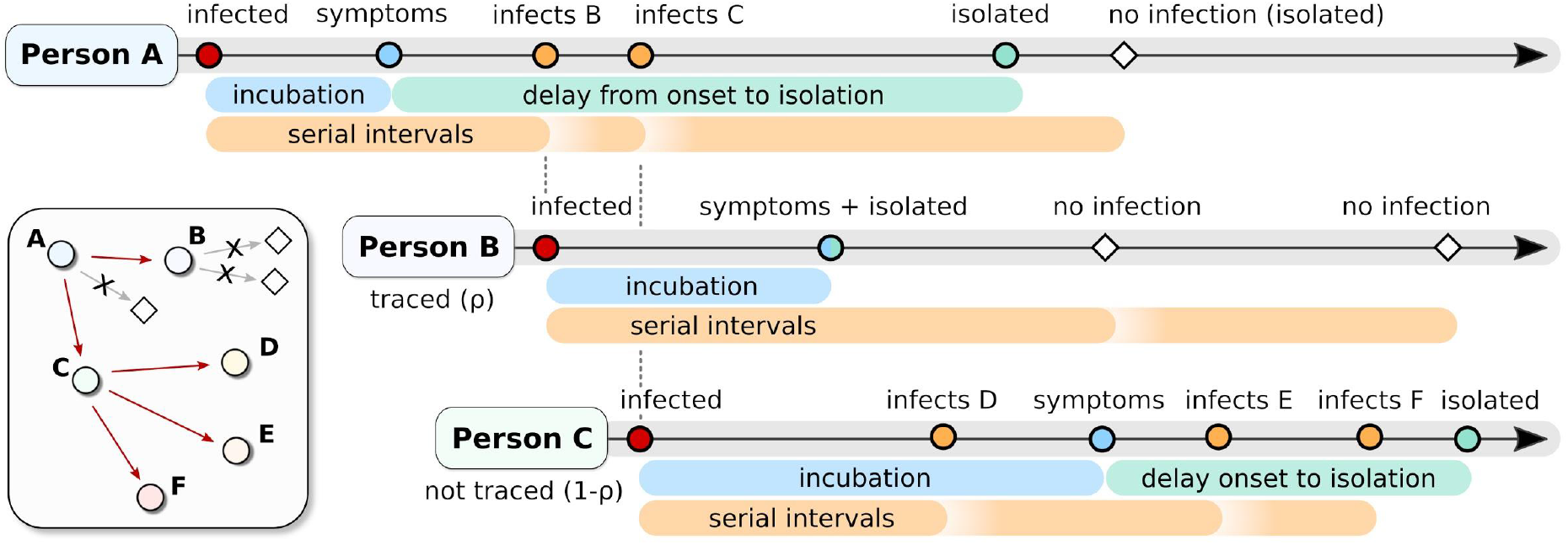
Example of the simulated process that starts with person A being infected. After an incubation period (blue) person A shows symptoms and is isolated at a time drawn from the delay distribution (green) (Table 1). A draw from the negative binomial distribution with mean *R*_*0*_ and distribution parameter determines how many people person A potentially infects. For each of those, a serial interval is drawn (orange). Two of these exposures occur before the time that person A is isolated. With probability *ρ*, each contact is traced, with probability *1-ρ* they are missed by contact tracing. Person B is successfully traced, which means that they will be isolated without a delay when they develop symptoms. hey could, however, still infect others before they are isolated. Person C is missed by contact tracing. This means that they are only detected if and when symptomatic, and are isolated after a delay from symptom onset. Because person C was not traced they infected two more people (E and F) in addition to person D than if they had been isolated at symptom onset. A version with asymptomatic transmission is given in Figure S8.

We initialised the branching process with 5, 20, or 40 cases to represent a newly detected outbreak of varying size. Initial symptomatic cases were then isolated after symptom onset with a delay drawn from the onset-to-isolation distribution (Table 1). Isolation was assumed to be 100% effective at preventing further transmission; therefore, in the model, failure to control the outbreak resulted from the lack of complete contact tracing and the delays in isolating cases rather than the inability of isolation to prevent further transmission. Either 100% or 90% of cases became symptomatic, and all symptomatic cases were eventually reported.

**Table 1.**
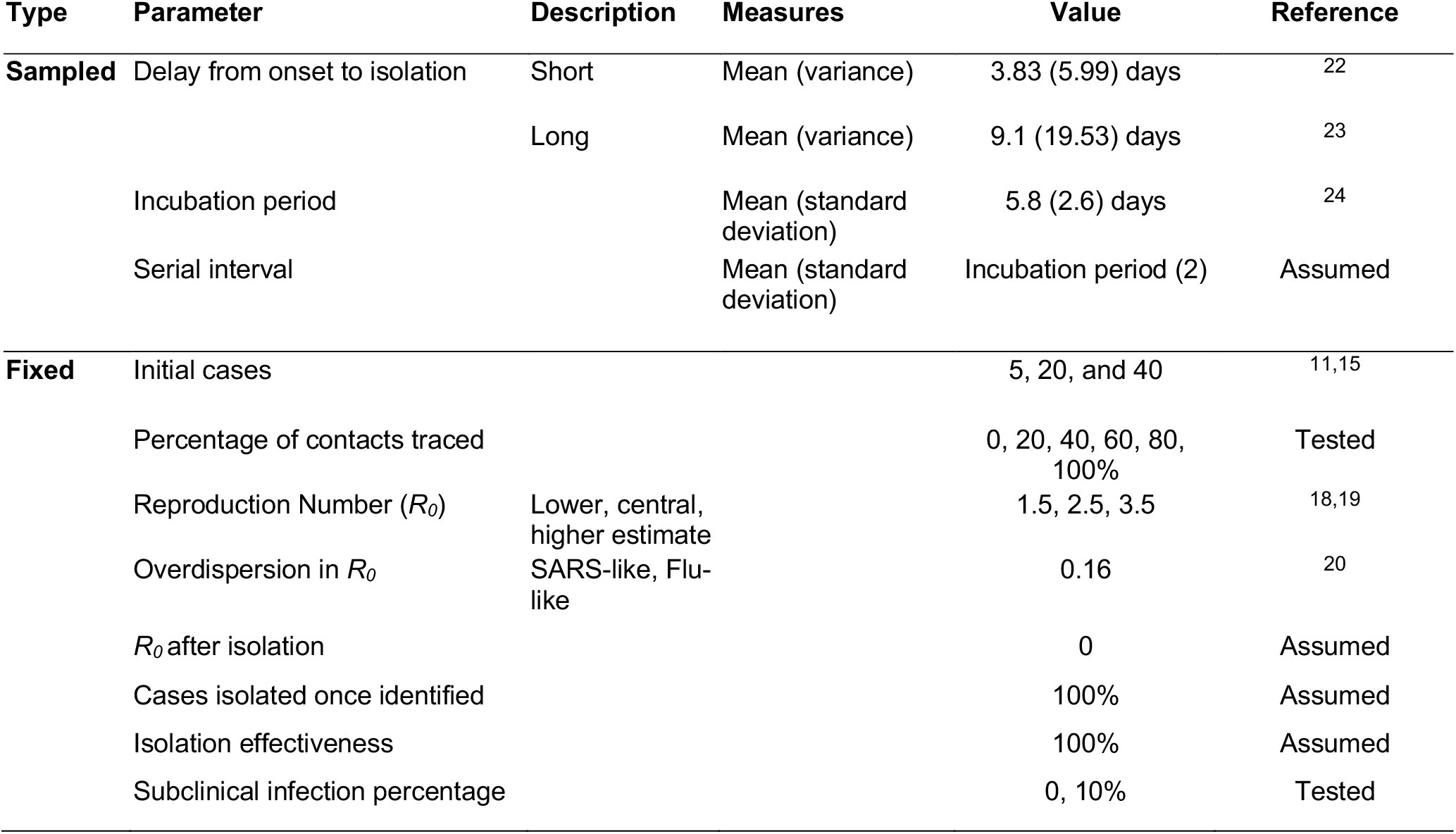
Parameter values for the model. Sampled values are probabilistically sampled during the simulation, and fixed values remain constant during the simulation.

| Type | Parameter | Description | Measures | Value | Reference |
| --- | --- | --- | --- | --- | --- |
| Sampled | Delay from onset to isolation | Short | Mean (variance) | 3.83 (5.99) days | 22 |
|  |  | Long | Mean (variance) | 9.1 (19.53) days | 23 |
|  | Incubation period |  | Mean (standard deviation) | 5.8 (2.6) days | 24 |
|  | Serial interval |  | Mean (standard deviation) | Incubation period (2) | Assumed |
| Fixed | Initial cases |  |  | 5, 20, and 40 | 11,15 |
|  | Percentage of contacts traced |  |  | 0, 20, 40, 60, 80, 100% | Tested |
| | Reproduction Number ($R_0$) | Lower, central, higher estimate | | 1.5, 2.5, 3.5 | 18,19 |
| | Overdispersion in $R_0$ | SARS-like, Flu-like | | 0.16 | 20 |
| | $R_0$ after isolation | | | 0 | Assumed |
|  | Cases isolated once identified |  |  | 100% | Assumed |
|  | Isolation effectiveness |  |  | 100% | Assumed |
|  | Subclinical infection percentage |  |  | 0, 10% | Tested |

Each newly infected case was identified through contact tracing with probability *ρ*. Secondary cases that had been traced were isolated immediately upon becoming symptomatic. Cases that were missed by contact tracing (probability 1- *ρ*) were isolated when they became symptomatic with a delay drawn from the onset-to-isolation distribution.

In addition, each case had an independent probability of being subclinical (asymptomatic), and were therefore not detected either by self report or if traced by contact tracing. New secondary cases caused by an asymptomatic case were missed by contact tracing and could only be isolated based on symptoms. The model includes isolation of symptomatic individuals only, i.e. no quarantine, so isolation cannot prevent transmission before symptom onset. Quarantining contacts of cases requires a considerable investment in public health resources, and has not been widely implemented for all contacts of cases^3,21^.

### Transmission scenarios

We ran 1,000 simulations of each combination of the proportion of transmission before symptom onset, *R*_*0*_, onset-to-isolation delay, the number of initial cases, and the probability that a contact was traced (Table 1).

We explored two scenarios of delay between symptom onset and isolation: “short” and “long” (Figure 2). The short delay was estimated during the late stages of the 2003 SARS outbreak in Singapore^20^ and the long delay was an empirical distribution calculated from the early phase of the 2019-nCoV outbreak in Wuhan^21^. We varied the percentage of contacts traced from 0-100% at 20% intervals to quantify the effectiveness of contact tracing.

**Figure 2:**
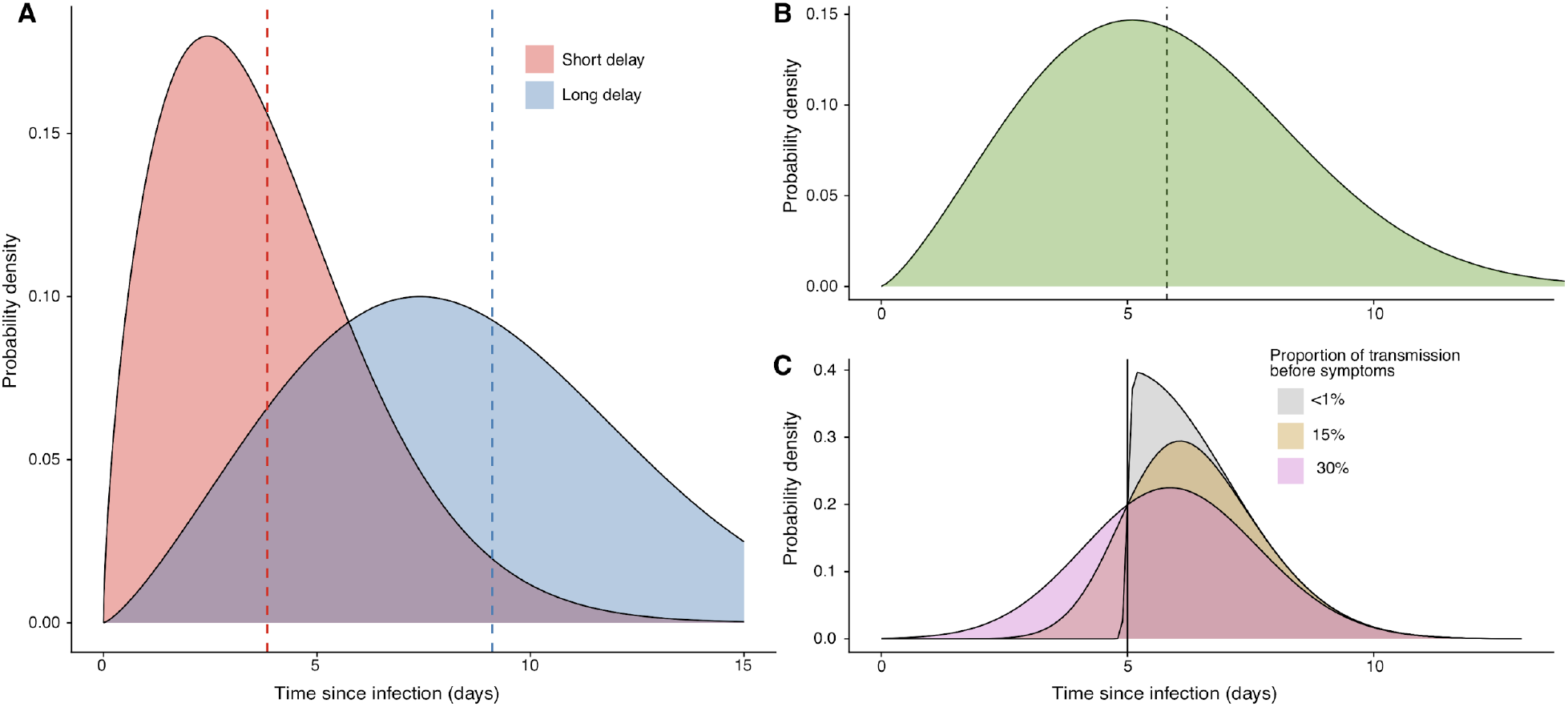
A) The short and long delay distributions between the onset of symptoms and isolation (mean marked by line). Parameter values and references are given in Table 1. B) The incubation distribution estimate fitted to data from the Wuhan outbreak by Backer *et al*.^24^. C) An example of the method used to sample the serial interval for a case that has an incubation period of 5 days. Each case has an incubation period drawn from the distribution in B, their serial interval is then drawn from a skewed normal distribution with the mean set to the incubation period of the case. In C, the incubation period was 5 days. The skew parameter of the skewed normal distribution controls the proportion of transmission that occurs before symptom onset, the three scenarios explored are <1% of transmission before onset (grey), 15% of transmission before onset (gold), and 30% of transmission before onset (pink).

The incubation period for each case was drawn from a Weibull distribution. A corresponding serial interval for each case was then drawn from a skewed normal distribution with the mean parameter of the distribution set to the incubation period for that case, a standard deviation of 2, and a skew parameter chosen such that a set proportion of serial intervals were shorter than the incubation period (meaning that a set proportion of transmission happened before symptom onset) (Figure 2). This sampling approach ensured that the serial interval and incubation period for each case was correlated, and prevents biologically implausible scenarios where a case can develop symptoms very soon after exposure but not become infectious until very late after exposure and *vice versa*.

There are many estimates of the reproduction number for the early phase of the 2019-nCoV outbreak in Wuhan, China^15,18,19,23,26,26–30^, and therefore we used the values 1.5, 2.5, and 3.5, which span most of the range of current estimates (Table 1). We used the secondary case distribution from the 2003 SARS outbreak^20^, and tested the effect of lower heterogeneity in the number of secondary cases^25^ as a sensitivity analysis (see supplement). We calculated the effective reproduction number (*R*_*eff*_) of the simulation as the average number of secondary cases produced by each infected person in the presence of isolation and contact tracing. We present results in relation to the baseline scenario of *R*_*0*_ of 2.5, 20 initial cases, a short delay to isolation, 15% of transmission before symptom onset, and 0% subclinical infection.

### Definition of outbreak control

Outbreak control was defined as no new infections between 12 and 16 weeks after the initial cases. Outbreaks that reached 5,000 cumulative cases were assumed to be too large to control within 12-16 weeks, and were categorised as uncontrolled outbreaks. Based on this definition, we reported the probability that an outbreak of a 2019-nCoV-like pathogen would be controlled within 12 weeks for each scenario, assuming that the basic reproduction number remained constant and no other interventions were implemented.

The probability that an outbreak is controlled gives a one-dimensional understanding of the difficulty involved in achieving control, because the model places no constraints on the number of cases and contacts that can be traced and isolated. In reality, the feasibility of contact tracing and isolation is likely to be determined both by the probability of achieving control, and the resources needed to trace and isolate infected cases^31^. We therefore reported the weekly maximum number of cases undergoing contact tracing and isolation for each scenario that results in outbreak control. Once the weekly number of cases reaches a certain point, it can overwhelm the contact tracing system and affect the quality of the contact tracing effort^32^.

### Availability of methods

All code is available as an R package (https://github.com/epiforecasts/ringbp).

## Results

### Effect of reproduction number on outbreak control

To achieve 90% of outbreaks controlled required 80% of contacts to be traced and isolated for scenarios with a reproduction number of 2.5 (Figure 3a). The probability of control was higher at all levels of contact tracing when the reproduction number was lower, and fell rapidly for a reproduction number of 3.5. At a reproduction number of 1.5, the effect of isolation is coupled with the chance of stochastic extinction resulting from overdispersion^20^, which is why some outbreaks were controlled even at 0% contacts traced.

**Figure 3:**
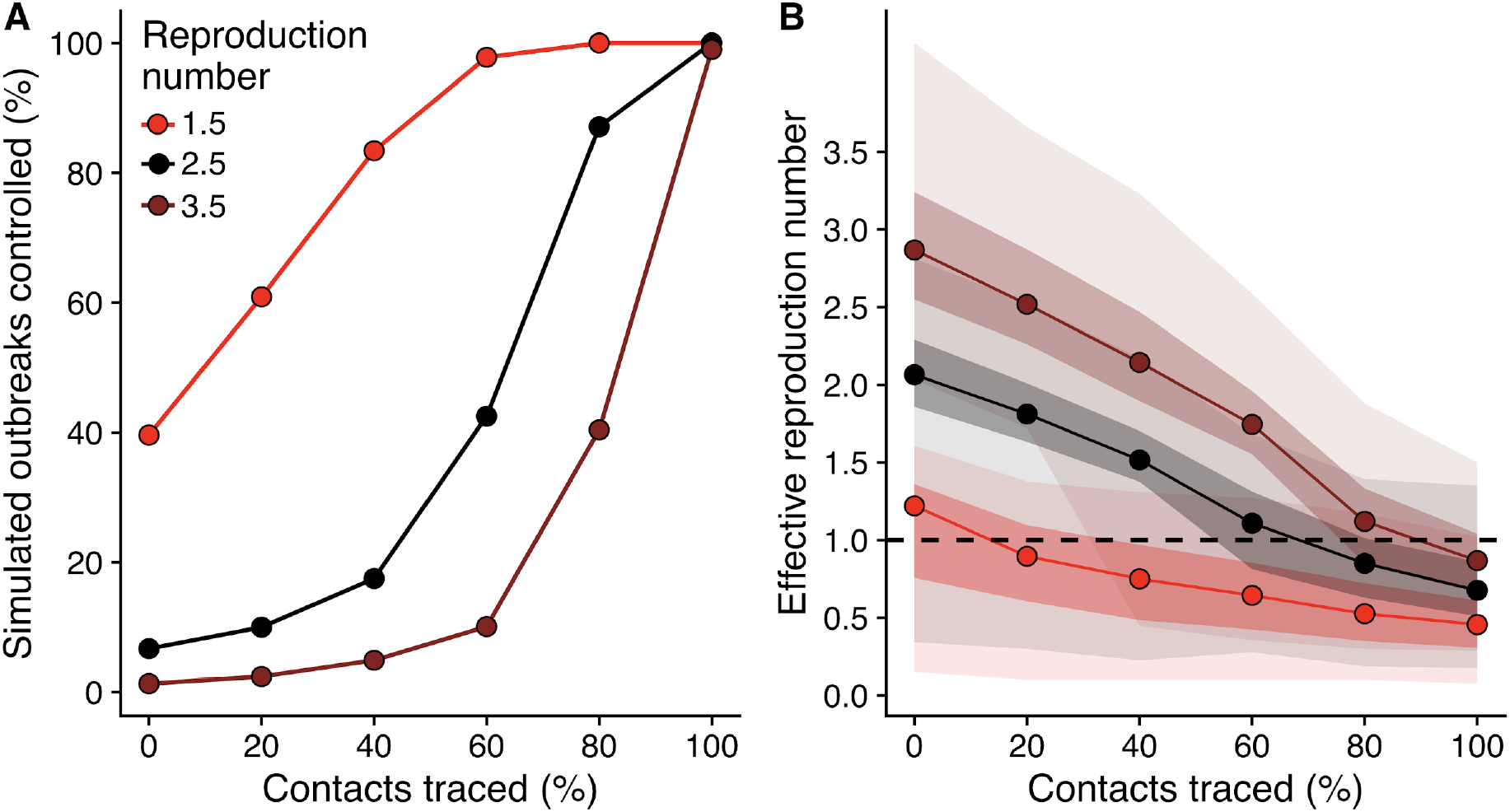
A) The percentage of outbreaks that are controlled for scenarios with varying reproduction number (*R*_*0*_), at each value of contacts traced. The baseline scenario is *R*_*0*_ of 2.5, 20 initial cases, a short delay to isolation, 15% of transmission before symptom onset, and 0% subclinical infection (black line). A simulated outbreak is defined as controlled if there are no cases between weeks 12 and 16 after the initial cases. See supplement for other scenarios. B) Effective reproduction number in the presence of case isolation and contact tracing. Median (line), and 50% and 95% intervals (shaded regions) are shown.

Isolation and contact tracing decreased transmission, as shown by a decrease in the effective reproduction number (Figure 3b). For the scenario where the basic reproduction number was 1.5, the median estimate rapidly fell below 1, which indicates that control is likely. For the higher transmission scenarios a higher level of contact tracing was needed to bring the median effective reproduction number below 1.

### Impact of transmission characteristics on probability of achieving control

The number of initial cases had a large impact on the probability of achieving control. With five initial cases, there was a greater than 50% chance of achieving control in 3 months, even at modest contact tracing levels (Figure 4a). More than 40% of these outbreaks were controlled with no contact tracing due to the combined effects of isolation of symptomatic cases and stochastic extinction. The probability of control dropped as the number of initial cases increased, and for 40 initial cases, even 80% contact tracing did not lead to 80% of simulations controlled within 3 months.

**Figure 4:**
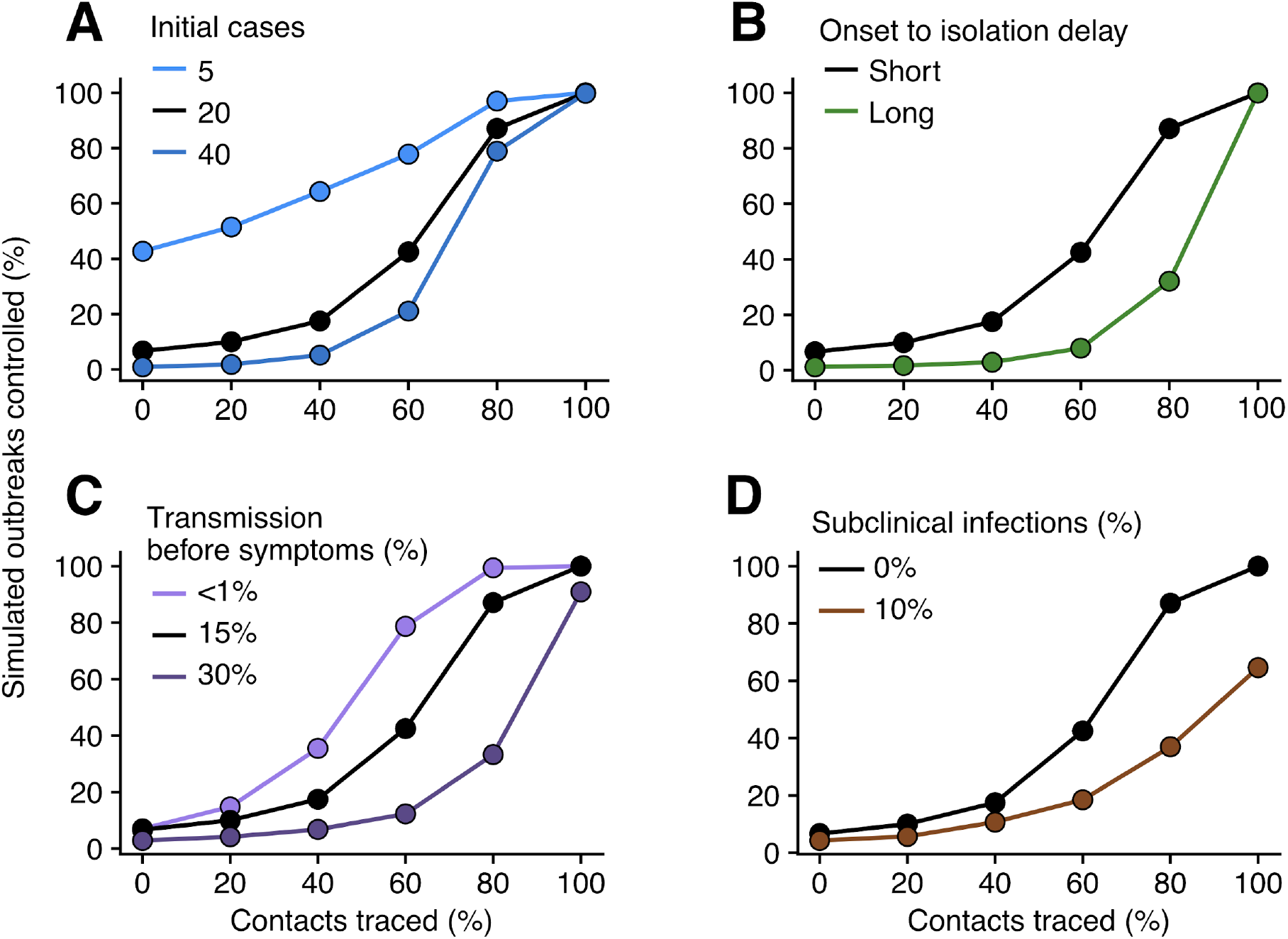
The percentage of outbreaks controlled for the baseline scenario (black), and varied number of initial cases (A), time from onset to isolation (B), percentage of transmission before symptoms (C), and proportion of subclinical (asymptomatic) cases (D). The baseline scenario is *R*_*0*_ of 2.5, 20 initial cases, a short delay to isolation, 15% of transmission before symptom onset, and 0% subclinical infection. Results for *R*_*0*_ = 1.5 and 3.5 are given in the supplement. A simulated outbreak is defined as controlled if there are no cases between weeks 12 and 16 after the initial cases.

The delay from symptom onset to isolation played a major role in achieving control of outbreaks (Figure 4b). At 80% of contacts traced, the probability of achieving control falls from 89% to 31% when there is a longer delay from onset to isolation. If there is no transmission before symptom onset then the probability of achieving control is higher for all values of contacts traced (Figure 4c). The difference between 15% and 30% of transmission before symptoms had a marked effect on probability to control. We found this effect in all scenarios tested (supplementary Figure S4). Including only 10% of cases being asymptomatic resulted in a decreased probability that simulations were controlled by isolation and contact tracing for all values of contact tracing (Figure 4d). For 80% of contacts traced, only 37% of outbreaks were controlled, compared with 89% without subclinical infection.

### Feasibility of contact tracing

In many scenarios there were between 25 and 100 symptomatic cases within a week (Figure 5), all of whom would need isolation and would require contact tracing. The maximum number of weekly cases may appear counterintuitive because a lower maximum number of weekly cases is not associated with higher outbreak control. This occurs because with better contact tracing it becomes possible to control outbreaks with higher numbers of weekly cases. The maximum number of weekly cases is lower if the initial number of cases is 5 and higher if it is 40 (see supplement). In the 2014 Ebola epidemic in Liberia, each case reported between 6 and 20 contacts^8^, and the number of contacts may be higher, as seen in MERS outbreaks^10^. Tracing 20 contacts per case could mean up to 2,000 contacts in the week of peak incidence.

**Figure 5:**
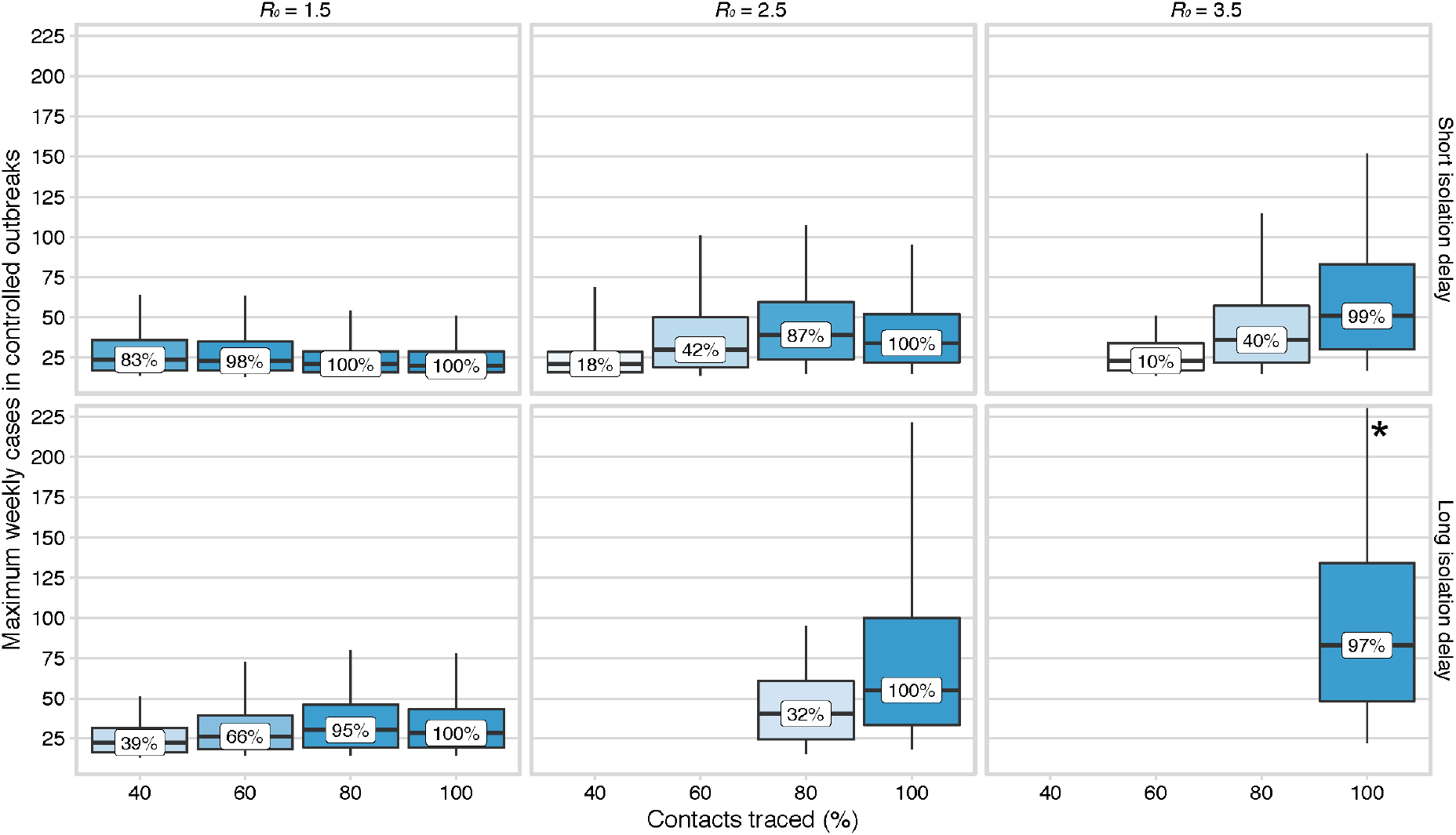
The maximum weekly cases requiring contact tracing and isolation in scenarios with 20 index cases that achieved control within 3 months. Scenarios vary by reproduction number and the mean delay from onset to isolation. 15% of transmission occurred before symptom onset, and 0% subclinical infection. The percentage of simulations that achieved control is shown in the boxplot. This illustrates the potential size of the eventually controlled simulated outbreaks, which would need to be managed through contact tracing and isolation. * indicates that the 95% interval extends out of the plotting region.

## Discussion

We determined conditions where case isolation, contact tracing, and preventing transmission by infected contacts would be sufficient to control a new 2019-nCoV outbreak in the absence of other control measures. We found that in many plausible scenarios, case isolation alone would be unlikely to control transmission within three months. Case isolation was more effective when there was little transmission before symptom onset and when the delay from symptom onset to isolation was shorter. Preventing transmission by tracing and isolating a larger proportion of contacts, thereby decreasing the effective reproduction number, improved the number of scenarios where control was likely to be achieved. However, these outbreaks required a large number of cases to be contact traced and isolated each week, which is of crucial concern when assessing the feasibility of this strategy. Subclinical infection markedly decreased the probability of controlling outbreaks within 3 months.

In scenarios where the reproduction number was 2.5, 15% of transmission occurred before symptom onset, and there was a short delay to isolation, at least 80% of infected contacts needed to be traced and isolated to give a probability of control of 90% or more. This echoes other suggestions that highly effective contact tracing will be necessary to control outbreaks in other countries^17^. In scenarios where the delay from onset to isolation was larger, similar to the delays seen in the early phase of the outbreak in Wuhan, the same contact tracing success rate of 80% achieved less than 40% probability of containing an outbreak. Higher pre-symptomatic transmission decreases the probability that the outbreaks were controlled, under all reproduction numbers and isolation delay distributions tested.

Our model does not include other control measures that may decrease the reproduction number and therefore also increase the probability of achieving control of an outbreak. At the same time, it assumes that isolation of cases and contacts is completely effective, and that all symptomatic cases are eventually reported. Relaxing these assumptions would decrease the probability that control is achieved. We also make the assumption that contact is required for transmission between two individuals, whereas transmission via fomites may be possible. This would make effective contact tracing challenging, and good respiratory and hand hygiene would be critical to reduce this route of transmission, coupled with environmental decontamination in healthcare settings.

We intentionally simplified our model to determine the effect of contact tracing and isolation on the control of outbreaks under different scenarios of transmission. However, as more data becomes available, the model can be updated, or tailored to particular public health contexts. It is likely that the robustness of control measures is affected both by differences in transmission between countries but also by the concurrent number of cases that require contact tracing in each scenario. Practically, there is likely to be an upper bound on the number of cases that can be traced, and case isolation is likely to be imperfect^33^. We reported the maximum number of weekly cases during controlled outbreaks but the capacity of response efforts may vary.

We explored a range of scenarios informed by the latest evidence on transmission of 2019-nCoV. Similar analyses using branching models have already been used to analyse the Wuhan outbreak to find plausible ranges for the initial exposure event size and the basic reproduction number^15,19^. Our analysis expands on this by including infectiousness before the onset of symptoms, case isolation, explicit modelling of case incubation periods and time to infectiousness. A key area of uncertainty is if and for how long individuals are infectious before symptom onset, and if asymptomatic or subclinical infection occurs. Both are likely to make the outbreak harder to control^24^.

The model could be modified to include some transmission after isolation (such as in hospitals) which would decrease the probability of achieving control. In addition, we define an outbreak as controlled if it reaches extinction by 3 months, regardless of outbreak size or number of weekly cases. This definition may be narrowed where the goal is to keep the overall caseload of the outbreak low. This may be of concern to both local authorities for reducing the healthcare surges, and may provide a way to limit geographic spread.

Our study indicates that in most plausible outbreak scenarios case isolation and contact tracing alone is insufficient to control outbreaks, and that in certain scenarios even near perfect contact tracing will still be insufficient, and therefore further interventions would be required to achieve control. However, rapid and effective contact tracing can also reduce the initial number of cases, which would make the outbreak easier to control overall. Effective contact tracing and isolation could contribute to reducing the overall size of an outbreak or bringing it under control over a longer time period.

### Contributors

RME conceived the study. JH, AG, SA, WJE, SF, RME designed the model. CIJ, TWR, NIB worked on statistical aspects of the study. JH, AG, SA, NIB programmed the model, and with RME, made the figures. AJK and JDM consulted on the code. All authors interpreted the results, contributed to writing the manuscript and approved the final version for submission.

## Data Availability

No data are used. Model code is available.

## Acknowledgements

The named authors (JH, SA, AG, NIB, CIJ, TWR, JDM, AJK, WJE, SF, RME) had the following sources of funding: JH, SA, JDM and SF were funded by the Wellcome Trust (grant number: 210758/Z/18/Z), AG and CIJ were funded by the Global Challenges Research Fund (grant number: ES/P010873/1), TWR and AJK were funded by the Wellcome Trust (grant number: 206250/Z/17/Z), and RME was funded by HDR UK (grant number: MR/S003975/1). This research was partly funded by the National Institute for Health Research (NIHR) (16/137/109) using UK aid from the UK Government to support global health research. The views expressed in this publication are those of the author(s) and not necessarily those of the NIHR or the UK Department of Health and Social Care. This research was partly funded by the Bill & Melinda Gates Foundation (INV-003174). This research was also partly funded by the Global Challenges Research Fund (GCRF) project ‘RECAP’ managed through RCUK and ESRC (ES/P010873/1).

We would like to acknowledge (in a randomised order) the other members of the LSHTM 2019-nCoV modelling group, who contributed to this work: Stefan Flasche, Mark Jit, Nicholas Davies, Sam Clifford, Billy J Quilty, Yang Liu, Charlie Diamond, Petra Klepac and Hamish Gibbs. Their funding sources are as follows: SF and SC (Sir Henry Dale Fellowship (grant number: 208812/Z/17/Z)), MJ, YL, PK (BMGF (grant number: INV-003174)), ND (NIHR (grant number: HPRU-2012-10096)), BJQ (grant number: NIHR (16/137/109)), CD & YL (NIHR (grant number: 16/137/109)), and HG (Department of Health and Social Care (grant number: ITCRZ 03010))

## Data sharing

No data were used in this study. The R code for the work is available at https://github.com/epiforecasts/ringbp.

